# Redundancy, quality appraisal, and discordance in the results of systematic reviews of early mobilization of critically ill adults. A meta-research protocol

**DOI:** 10.1101/2023.04.05.23288203

**Authors:** Ruvistay Gutierrez-Arias, Dawid Pieper, Peter Nydahl, Felipe González-Seguel, Yorschua Jalil, Maria-Jose Oliveros, Rodrigo Torres-Castro, Pamela Seron

## Abstract

**Introduction:** In adult patients in intensive care units (ICU), early mobilization is one of the central non-pharmacological interventions studied for recovery from critical illness. Several systematic reviews (SRs) have been conducted to determine the effect of this intervention on ICU-acquired weakness (ICU-AW) with heterogeneous methodology and results. Redundancy in conducting SRs, unclear justification when leading new SRs or updating, and discordant results of SRs on the same research question may be generating research waste that makes it difficult for clinicians to keep up to date with the best available evidence. Therefore, this meta-research aims to assess the redundancy, methodological and reporting quality, and potential reasons for discordance in the results reported by SRs conducted to determine the effectiveness of early mobilization in critically ill adult patients on different clinical outcomes.

**Methods:** A meta-research of early mobilization SRs in critically ill adult patients will be conducted. A sensitive search of MEDLINE (Ovid), Embase (Ovid), CINAHL (EBSCOhost), Cochrane Library, Epistemonikos, and other search resources will be conducted. Two independent reviewers will perform study selection, data extraction, and quality appraisal. Discrepancies will be resolved by consensus or a third reviewer. The redundancy of SRs will be assessed by the degree of overlap of primary studies. In addition, the justification for conducting new SRs will be evaluated with the “Evidence-Based Research” framework. The methodological quality of the SRs will be assessed with the AMSTAR 2 tool and the quality of the reports through compliance with the PRISMA statement. To assess the potential reasons for discordance in the results of the SRs, only SRs that an MA has carried out will be analyzed, considering divergence in results and their interpretation.

**Expected results:** The analysis of this meta-research will assess the redundancy in the conducting of SR on the mobilization of critically ill adult patients, their methodological quality, and the quality of the reporting of their findings, as well as the causes of possible discrepancies between their results. These findings could guide the development of better and more timely SRs on the effectiveness of early mobilization of adult critically ill patients. The decrease in waste research could facilitate evidence-based decision-making by stakeholders.

**Registration number:** osf.io/kxwq9

## INTRODUCTION

Critically ill adult patients may present with various complications from hospitalization in an intensive care unit (ICU) stay [1,2]. Being on mechanical ventilation, sedation, neuromuscular blockade, and the mobility restrictions imposed by the context of critical illness, as well as barriers derived from invasive devices in critically ill patients [3–5], create an environment that can facilitate cognitive [6] and neuromusculoskeletal complications [7], among others.

One of the main ones is ICU-acquired weakness (ICU-AW) [8,9]. The prevalence of ICU-AW is variable [10]; however, it is a problem that should be considered a priority in managing critically ill patients. ICU-AW is associated with other structural and functional impairments that may lead to patient activities and participation restrictions. Decreased quality of life, reduced participation in social activities, and low frequency of return-to-work activities in the post-discharge setting have been reported [11–13].

This health condition typically appears generalized and symmetrical, affecting limb and respiratory muscles. This weakness may be due to altered nerve stimulus conduction (critical illness polyneuropathy), altered muscle contraction due to myogenic disturbance (critical illness myopathy), or a mixture of both pathophysiological processes (critical illness neuromyopathy) [14,15]. The diagnosis of ICU-AW can be performed in different ways [16]. The most used in clinical practice is the manual assessment of muscle strength of the four limbs using the Medical Research Council sum score scale (MRC-SS) [17].

Early mobilization is one of the central non-pharmacological interventions studied to prevent or recover from ICU-AW. While the definition of early mobilization is not agreed upon [18], it is expected that this intervention should be applied as early as possible to critically ill patients, starting with passive mobilization of limbs and other body segments, continuing with active mobilization as early as possible, and with functional transitional exercises to higher positions including assisted ambulation. In addition, devices to support passive and active mobilization, such as cycles or cycle ergometers, can be added [19].

Positive effects on muscle strength, length of ICU and hospital stay, and duration of mechanical ventilation, among others, have been reported [20–23]. However, the evidence from primary studies on the effectiveness of early mobilization is inconsistent. Therefore, several systematic reviews (SRs) have been conducted to determine the effect of this intervention trough different modalities, such as assessment of the quality or risk of bias of the primary studies and pooled data analysis (meta-analysis (MA)).

SRs are considered to have the highest level of evidence to establish the effectiveness and safety of any intervention in different health conditions [24]. This type of secondary study is the basis for developing recommendations in clinical practice guidelines [25]. However, the number of SRs published recently has increased exponentially [26], and some SRs seeks to answer the same research question, finding limited methodological quality among them.

Redundancy in SRs [27], the unclear justification provided when conducting a new SR or updating a previous one [28], and the discordant results of SRs on the same research question may lead to difficulties for clinicians to keep up to date and identify the best available evidence [29–31]. Therefore, this meta-research aims to assess the redundancy, methodological and reporting quality, and potential reasons for discordance in the results reported by SRs conducted to determine the effectiveness of early mobilization in critically ill adult patients on different clinical outcomes. In addition, this meta-research aims to explore the reasons given by the authors of SRs when justifying the conduct of a new SR for the same research question, the use of previous SRs to guide the design of their studies, and whether the findings of their SRs are discussed based on previously published SRs.

## METHODS

There are no standard guidelines that can be used for meta-research studies. However, in many aspects our work will resemble an overview of SRs of interventions. Thus we will follow the recommendations proposed by the Cochrane Handbook for Systematic Reviews of Interventions where appropriate [32]. Furthermore, this protocol was reported according to Preferred Reporting Items for Systematic Review and Meta-Analysis Protocols (PRISMA-P) statement, where appropriate, [33] and was registered in Open Science Framework (OSF) under the number osf.io/kxwq9. The findings of this meta-research will be guided by the Preferred Reporting Items for Overviews of Reviews (PRIOR) statement [34].

The SRs identified in our overview on the effectiveness of physical rehabilitation interventions on neuromusculoskeletal function in critically ill patients will be considered the basis of this meta-research [35]. However, different eligibility criteria will be applied in terms of the population and intervention studied.

### Eligibility criteria

#### Type of studies

Intervention SRs, with or without meta-analysis, that have considered primary studies with a randomized (RCTs) or non-randomized clinical trial (non-RCTs) design will be included. SRs that perform only network meta-analyses without including pairwise comparative analyses of interventions (conventional meta-analyses) will be excluded.

Considering that there are different definitions of SRs [36], for this meta-research, intervention SRs will be defined as an evidence synthesis study that aims to answer pre-defined research questions using explicit, reproducible methods to identify, critically appraise and combine results of primary research studies aimed at determining the effectiveness of any intervention on different health conditions [37].

#### Type of participants

SRs that consider adult patients, with majority (>50%) being on invasive or non-invasive mechanically ventilation at least once during the stay on ICU, will be included. The illness or health condition that led to the need for critical care shall not limit inclusion.

In contrast to the eligibility criteria of our overview of SRs protocol, only the adult population will be considered because it is in this population that most SRs have been conducted.

#### Type of interventions

SRs that consider early mobilization as an intervention, as defined by the authors of the SRs, will be included. They may have but are not limited to the passive mobilization of limbs or another body segment [38,39], exercises involving active patient participation [40], and the use of assistive devices such as upper and lower extremity cycling or cycle ergometer [19,38,39].

#### Type of comparators

SRs that consider any intervention in the control groups of the primary studies will be included. These interventions may include usual care, placebo, sham, delayed mobilization, or other physical rehabilitation interventions.

#### Types of outcomes

SRs that have addressed the effectiveness of early mobilization on at least one of the following outcomes will be included:

- Mobility: Outcome that can be measured with any generic or specific scale to assess functionality in ICU, such as Functional Status Score for the Intensive Care Unit (FSS-ICU) [41], ICU mobility scale (IMS) [42], The Chelsea Critical Care Physical Assessment Tool (CPAx) [43], or any other measure to assess mobility.
- Muscle strength: Outcome that can be measured using a manual scale, for example, MRC-SS [44], or using a device that allows the assessment of handgrip strength [45] or the pressures generated by the respiratory muscles [46], among others.
- Muscle mass: Outcome which can be measured by muscle circumference measurement, ultrasonography, dual-energy X-ray absorptiometry, computed tomography scan [47], among other.
- Duration of mechanical ventilation: number of days patients remain on invasive ventilatory support.
- ICU length of stay: days between admission to the ICU and discharge to a less complex unit.
- Mortality: Due to any cause and which can be reported according to different follow-up points, for example, mortality in ICU, hospital, 90 days, 180 days, 360 days, the number of deaths due to a given cause.
- Incidence and duration of delirium: Outcome that can be measured with a scale such as the Confusion Assessment Method for the Intensive Care Unit (CAM-ICU) [48], among others.
- Unwanted safety events: Outcome that can be measured as the incidence of any unwanted safety events associated with the delivery of physical rehabilitation interventions reported by SRs.

### Search strategy

A systematic search with a sensitive approach will be conducted in different electronic databases and other search resources. MEDLINE (through Ovid), Embase (through Ovid), CINAHL (through EBSCOhost), Cochrane Library, and Epistemonikos will be searched using controlled language (i.e., MeSH, Emtree, and CINAHL Subject Headings) and key terms. In addition, the International Prospective Register of Systematic Reviews (PROSPERO), International Platform of Registered Systematic Review and Meta-analysis Protocols (INPLASY), and Open Science Framework (OSF) registries will be reviewed.

In addition, the references of the SRs included in this overview will be manually searched using the Citationchaser tool [49], and experts in critical patient rehabilitation will be consulted to identify potential SRs that meet the eligibility criteria of this overview.

The search strategy for MEDLINE (Ovid) (Table 1) was constructed following the Peer Review of Electronic Search Strategies (PRESS) statement [50]. The search strategy for MEDLINE (Ovid) was built following the PRESS statement, which will be adapted for the other electronic databases and search resources. The Canadian Agency for Drugs and Technologies in Health (CADTH) filter was used to identify studies with an SR design [51].

**Table 1.**
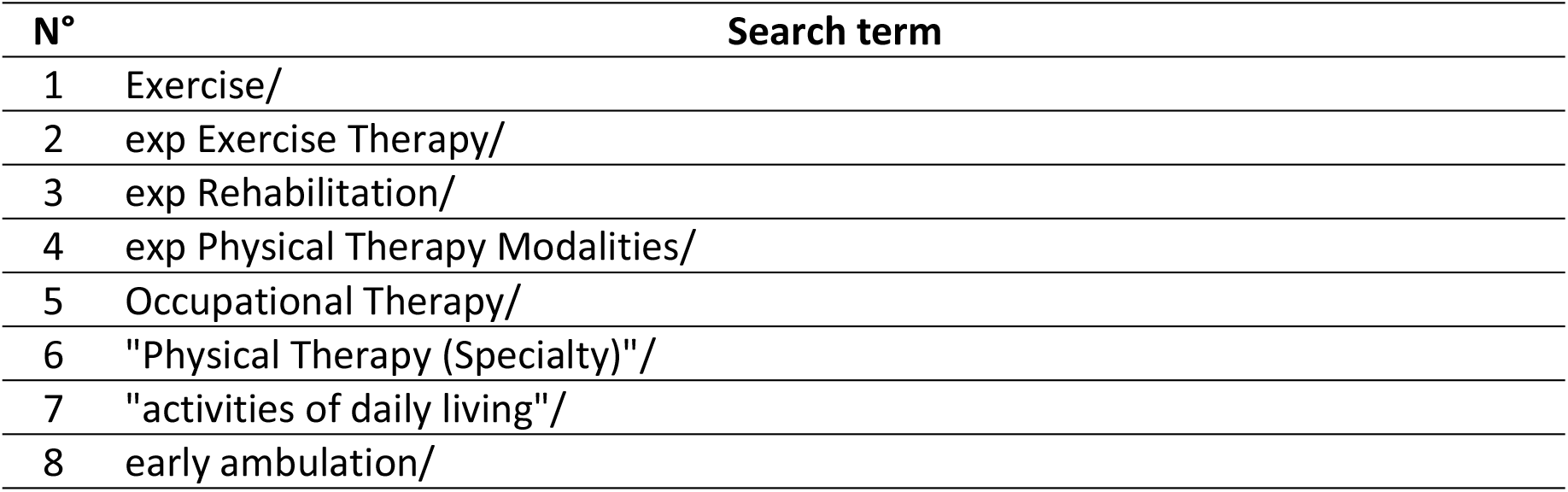
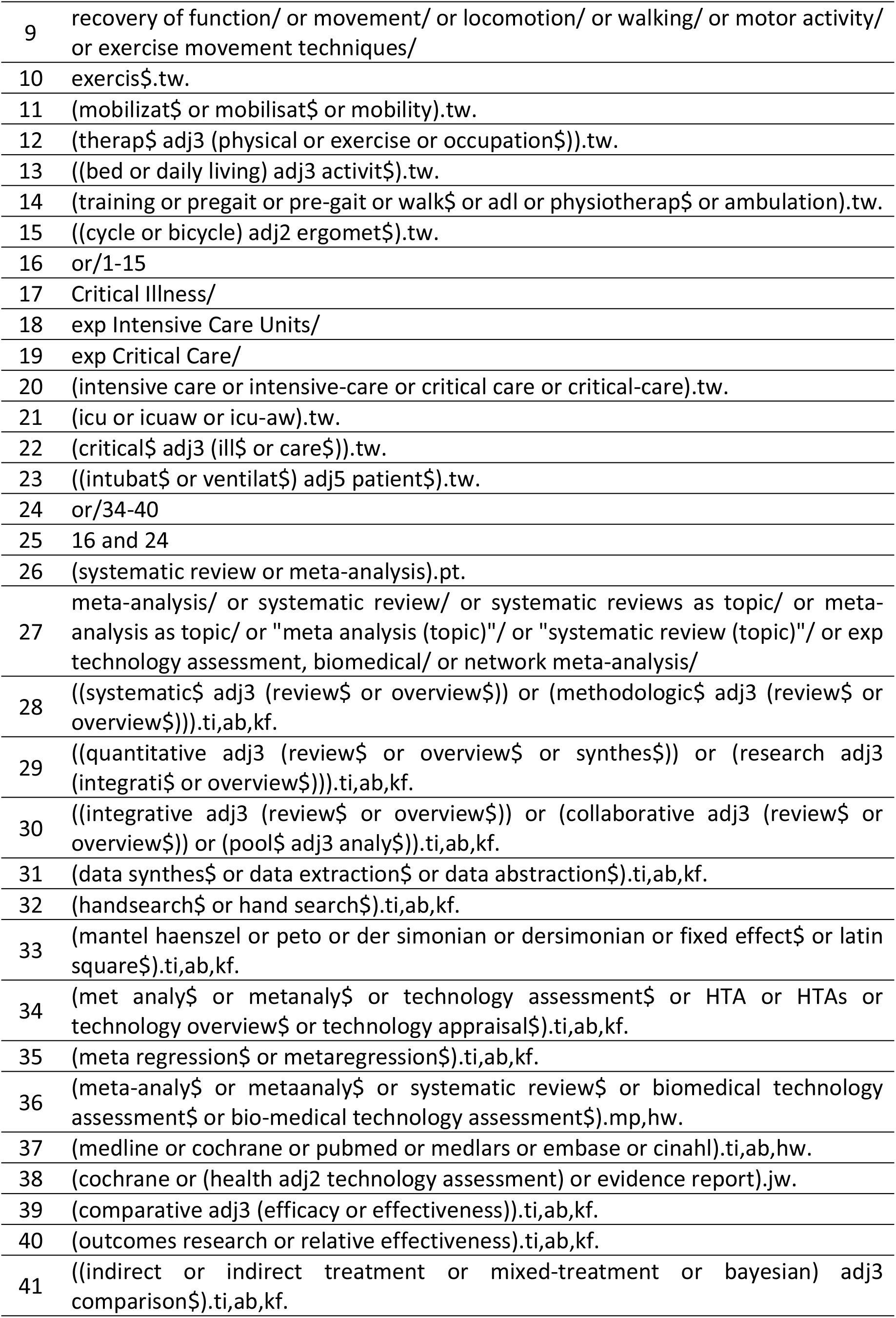
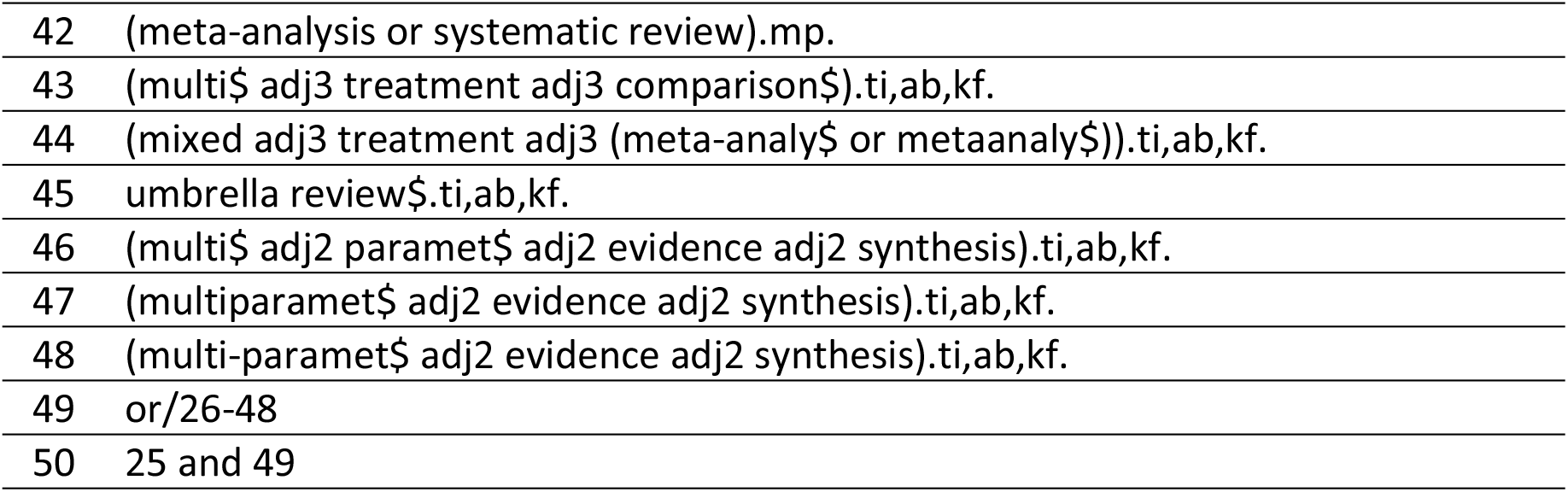
Search strategy for MEDLINE (Ovid).

### Study selection

Two reviewers will independently check records identified by the search strategy for compliance with the eligibility criteria. Irrelevant documents will be excluded by reading the title and abstract and then determining the inclusion of SRs by reading the full text. Disagreements will be resolved by consensus or by a third reviewer. The Rayyan® application will be used to improve the efficiency of this meta-research stage [52].

### Data extraction

Two reviewers will independently extract data from the SRs. An extraction form explicitly created for this study will be used, piloted with data extraction from 5 SRs, and then adapted according to the reviewers’ feedback in the piloting. This form will seek to extract data to describe the characteristics of the publication, general characteristics of the SRs, reported outcome data, quality or risk of bias of the primary studies included, and certainty of evidence (Table 2). In addition, the methodological and reporting quality of the SRs will be rated in the data extraction form. Disagreements will be resolved by consensus or by the involvement of a third reviewer.

**Table 2.** Data to extract.

| Domain | Data to extract |
| --- | --- |
| Bibliometric characteristics | a) Title<br>b) Authors<br>c) Countries involved in the SR<br>d) Year of publication<br>e) Journal<br>f) Journal impact factor at the time of publication of the SR<br>g) Protocol registration<br>h) Date of manuscript submission and publication |
| General characteristics of the SRs | a) Inclusion and exclusion criteria for primary studies according to the acronym PICO<br>b) Population description<br>c) Definition of early mobilization proposed by the SRs' authors<br>d) Electronic databases and other search resources considered by the SR<br>e) Search timeframe<br>f) Study designs included by the SR<br>g) Publication status<br>h) Reasons for exclusion of primary studies reviewed in full text<br>i) Previous early mobilization SRs cited in introduction*<br>j) Previous early mobilization SRs cited in discussion*<br>k) Qualitative or survey-based studies of the preferences or values of the end-users of the SRs cited in the introduction*<br>* Together with the sentences or paragraphs mentioned |
| Reported outcome data | a) Outcomes initially considered by SRs<br>b) Outcomes reported by SRs<br>c) Scales, questionnaires, or instruments used to assess different outcomes<br>d) Type of synthesis of results (meta-analysis or narrative)<br>e) Results data for each outcome reported |
| Quality or risk of bias of the primary studies | a) Instrument for assessing the methodological quality or risk of bias of included primary studies<br>b) Results of the assessment of the methodological quality or risk of bias of the included studies |
| Certainty of evidence | a) Instruments or framework used to assess the certainty of the evidence<br>b) Results of the assessment of the certainty of the evidence |
| Conclusion | a) Conclusions on the effectiveness of early mobilization<br>b) Recommendations for clinical practice<br>c) Recommendations for research |

The search strategy will not use language, date or publication status restrictions.

### Methodological appraisal

Two reviewers will independently assess the methodological quality of the SRs included in this overview using “A MeaSurement Tool to Assess systematic Reviews 2” (AMSTAR 2) [53]. Disagreements will be resolved by consensus or by the involvement of a third reviewer.

This tool includes 16 items and considers seven as critical:

1. Protocol registered before the commencement of the review;
2. Adequacy of the literature search;
3. Justification for excluding individual studies;
4. Risk of bias from individual studies being included in the review;
5. Appropriateness of meta-analytical methods;
6. Consideration of risk of bias when interpreting the results of the review;
7. Assessment of presence and likely impact of publication bias.

SRs will be classified according to the overall confidence in their results as High, Moderate, Low, and Critically Low, according to the following criteria:

- High: No or one non-critical weakness. The SR provides an accurate and comprehensive summary of the results of the available studies that address the question of interest.
- Moderate: More than one non-critical weakness. The SR has more than one weakness but no critical flaws. It may provide an accurate summary of the results of the available studies that were included in the review.
- Low: One critical flaw with or without non-critical weaknesses. The SR has a critical flaw and may not provide an accurate and comprehensive summary of the available studies that address the question of interest.
- Critically low. More than one critical flaw with or without non-critical weaknesses. The SR has more than one critical flaw and should not be relied on to provide and accurate and comprehensive summary of the available studies.

### Reporting quality

Two reviewers will independently assess SR authors’ adherence to the PRISMA statement when reporting their findings. Compliance will be assessed for the updated version [54]. Disagreements between the reviewers will be resolved by consensus or by a third reviewer.

### Data analysis and evidence synthesis

The SR selection process will be reported in narrative form with a PRISMA-type flow chart [54].

To assess the redundancy of SRs, a matrix will be created that cross-references the SRs identified by the search strategy with the primary studies included by these SRs. This will be done at the SR and outcome level. In addition, from these matrices, the corrected covered area (CCA) [55] will be calculated without considering any structural missing data and considering the chronological and primary study design structural missing data. The ccaR package (https://github.com/thdiakon/ccaR) will be used [56]. The crossover matrix of the SRs and primary studies included will be reported. In addition, heat map graphics will be presented to inform the degree of overlap of primary studies at the SR and outcome level.

In addition, the Evidence-Based Research framework will be used to assess whether, as new SRs were published, preceding SRs were cited or used to 1) justify the conduct of a new evidence synthesis study, 2) contribute to the design of new evidence synthesis studies, and 3) discuss the findings of new SRs considering preceding evidence synthesis studies [57–59]. For this purpose, five questionable research practices will be assessed through content analysis based on what is reported in the SRs’ articles (Table 3) [28].

**Table 3.**
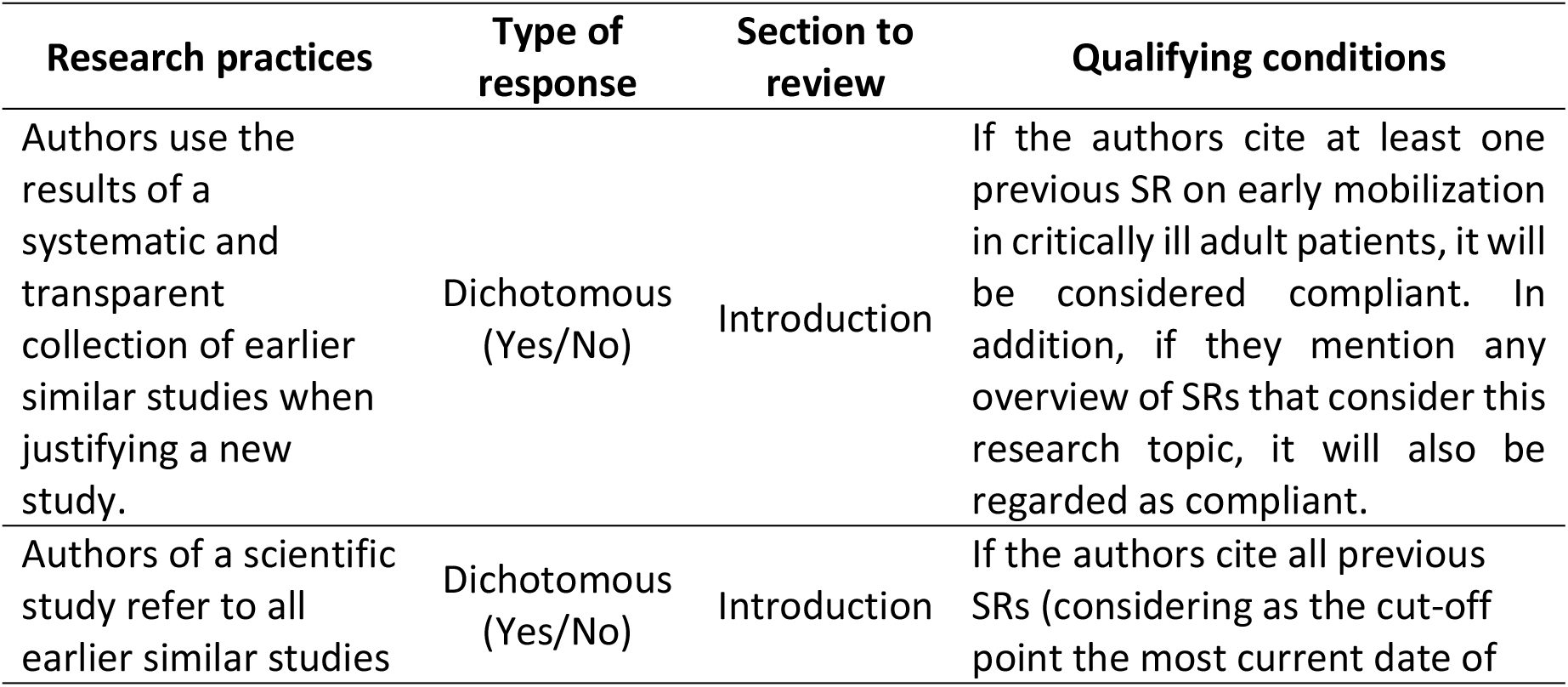
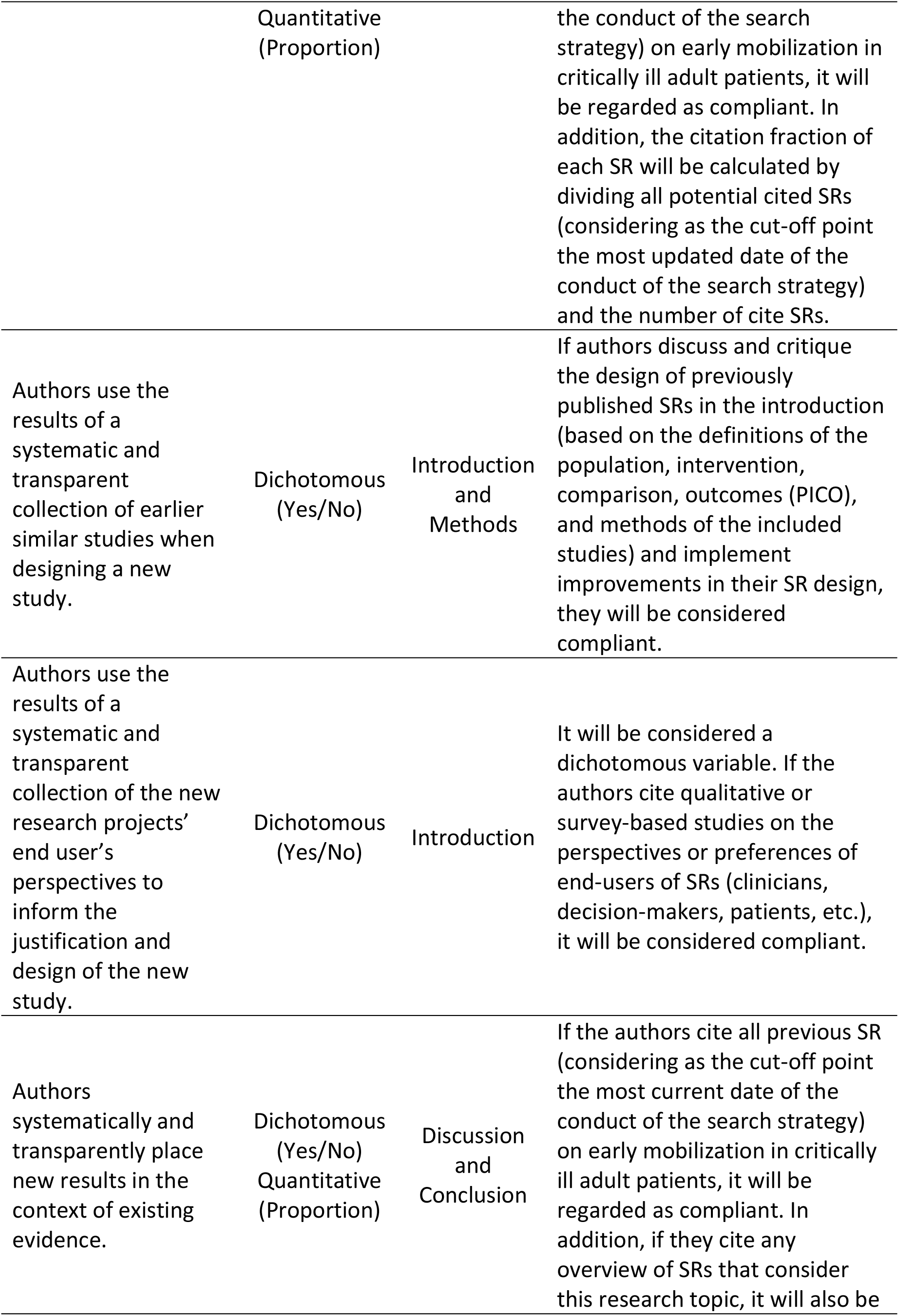
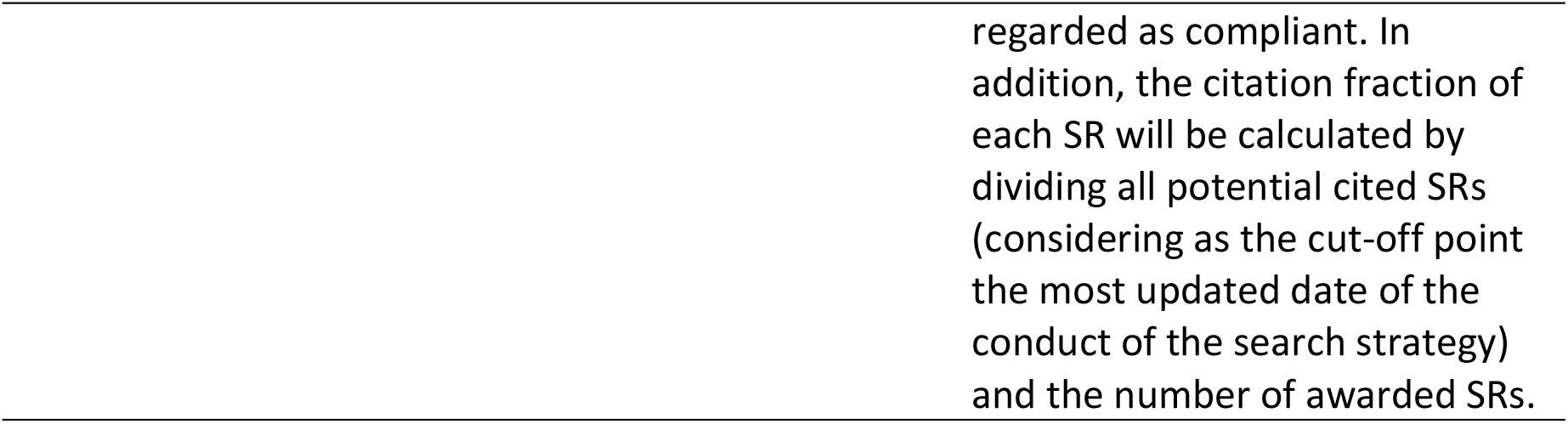
Evaluation of research practices in the Evidence-Based Research framework.

To assess the potential reasons for discordance in the results of the SRs, only SRs that have been carried out by an MA will be considered as a strategy to synthesize the results. The exploratory analysis will be conducted considering divergence in results or their interpretation:

#### Divergent results

A divergent result is defined as the variation between the SRs of the effect estimators’ values and their 95% CIs. The potential causes of variation to be explored will be:

a. Search date: the most recent search date reported by the SRs shall be considered.
b. Search resources: electronic databases and other search resources used by SRs will be considered.
c. Eligibility criteria: The definitions of eligibility criteria according to the PICO framework and the primary study designs included by the SRs will be considered.
d. Publication status: Consideration will be given to whether SRs included studies published only as abstracts in conference proceedings.
e. Excluded studies: reasons for exclusion of primary studies evaluated in full text will be considered.
f. Synthesis of outcome data: statistical methods for conducting meta-analyses (e.g., random effects vs. fixed effects), and data used to estimate the effect of the intervention (e.g., final scores vs. changes in scores from baseline) will be considered.

#### Divergent interpretations

Divergent interpretation shall be understood as variation in the conclusions in terms of the language used. This analysis will be performed by grouping SRs that determine that the effect estimator calculated using MA is 1) in favor of the intervention, 2) in favor of the comparator, and 3) neither in favor of the intervention nor the comparator. The potential causes of variation to be explored will be:

a. Risk of bias: the tool or scale used to assess the risk of bias of the included studies and the rating of the included studies will be considered.
b. Certainty of the evidence: consideration will be given to whether any framework was used to assess the certainty of the body of evidence (e.g., GRADE framework) and the grading of the evidence.
c. Statistical vs. clinical significance: we will consider whether the interpretation of the effectiveness of the studies was made based on statistical significance or by taking into account the minimally important clinical difference (clinical significance).
d. Conclusion: consideration will be given to whether the authors’ conclusions were made based on risk of bias or certainty of evidence.

## DISCUSSION

It is expected that the findings of this meta-research will make it possible to assess the redundancy in the conducting of SR on the mobilization of critically ill adult patients, their methodological quality, and the quality of the reporting of their findings, as well as the causes of possible discrepancies between their results.

The need to conduct new SRs that answer the same research question to be evaluated. The review of SR protocol registries [60] and implementation of the “evidence-based research” framework should be mandatory to reduce redundancy and research waste. This could allow for more efficient utilization of financial and human resources and make it easier for clinicians to keep up to date in their field.

## Data Availability

No datasets were generated or analysed during the current study. All relevant data from this study will be made available upon study completion.

## Ethics and dissemination

As meta-research, this study does not involve the participation of people whose rights may be violated. However, this overview will be developed rigorously and systematically to achieve valid and reliable results.

The findings of this meta-research study will be presented at conferences and published in a peer-reviewed journal related to rehabilitation, critical care, or research methodology.

## Notes

### Competing Interest Statement

The authors have declared no competing interest.

### Funding Statement

The author(s) received no specific funding for this work.

